# White Matter Abnormalities in Body Integrity Dysphoria

**DOI:** 10.1101/2021.08.18.21262224

**Authors:** Gianluca Saetta, Kathy Ruddy, Laura Zapparoli, Martina Gandola, Gerardo Salvato, Maurizio Sberna, Gabriella Bottini, Peter Brugger, Bigna Lenggenhager

## Abstract

“Body integrity dysphoria” (BID) is a severe condition affecting non-psychotic individuals where a limb may be experienced as not being part of the body, despite normal anatomical development and intact sensorimotor functions. Limb amputation is desired for restoring their own identity. We previously demonstrated altered brain structural (gray matter) and functional connectivity in 16 men with a long-lasting and exclusive desire for left leg amputation. Here we aimed to identify in the same sample altered patterns of white matter structural connectivity. Fractional anisotropy (FA), derived from Diffusion Tensor Imaging data, was considered as a measure of structural connectivity. Results showed reduced structural connectivity of: i) the right superior parietal lobule (rSPL) with the right cuneus, superior occipital and posterior cingulate gyri ii) the pars orbitalis of the right middle frontal gyrus (rMFGOrb) with the putamen iii) the left middle temporal gyrus (lMTG) with the pars triangularis of the left inferior frontal gyrus. Increased connectivity was observed between the right paracentral lobule (rPLC) and the right caudate nucleus. By using a complementary method of investigation, we confirmed and extended previous results showing alterations in areas tuned to the processing of the sensorimotor representations of the affected leg (rPCL), and to higher-order components of bodily representation such as the body image (rSPL). Alongside this network for bodily awareness, other networks such as the limbic (rMFGOrb) and the mirror (lMTG) systems showed structural alterations as well. These findings consolidate current understanding of the neural correlates of BID, which might in turn guide diagnostics and rehabilitative treatments.

## Introduction

“Body integrity dysphoria” (BID) represents a heterogeneous class of disorders characterized by dissatisfaction with the ordinal body morphology or functionality in non-psychotic individuals (Brugger et al., 2016). In its most typical form, one limb can be experienced as not being part of the body despite healthy anatomical development and intact sensorimotor functions (Stone et al., 2020). This feeling often leads to the desire for its amputation. While human beings commonly fear the idea of having a limb removed (Rozin et al., 2009), paradoxically, in BID individuals, the desired amputation would let them “*feel more complete”* (First, 2005) BID onset is typically in early childhood, or the desire has been lasting “for as long as the individual can remember” (Brugger et al., 2016). An exacerbation of the condition is sometimes seen at age 30-50 years, when the desire may be so distressing that the affected individuals might engage in hazardous behaviors like self-amputation (Sedda and Bottini, 2014). Individuals with BID also often engage in the mimicking of the status of an amputee by moving in a wheelchair, using crutches, or binding up the affected limb (i.e., pretending behavior) (First and Fisher, 2012), which can result in harmful consequences such as reduced blood supply to the affected limb. The prevalence of the disorder in the general population is unknown. However, although systematic epidemiological and clinical studies on BID are still lacking, BID has been found to be more common in men than in women and to predominately affect only one side of the body, typically the left lower limb (Brugger et al., 2016). BID individuals may also exhibit unusual erotic attraction to amputees lacking the limb, which is not felt to be part of their own body (Ramachandran et al., 2009). They may be sexually aroused by picturing themselves as an amputee (Blom et al., 2017). This behavioral feature has led to the initial conception of BID as a paraphilia (see *apotemnophilia;* Money et al., 1977). BID has also been referred to as a neurological syndrome (*xenomelia*; McGeoch et al., 2011), a personality disorder (First, 2005), or an internet-induced madness (Charland, 2004). Only very recently, in the release of the 11th Revision of the International Classification of Diseases (ICD 11), BID has been included in an official diagnostic manual, where it is listed among the developmental disorders and labelled a “disorder of bodily distress or bodily experience” (ICD-11). As specific diagnostic criteria and effective treatments still await to be identified, and the suffering of BID individuals is persistently high, there is a pressing need for research in this area. Indeed, to date, the psychotherapeutic treatments alone have proven ineffective or produced controversial effects on alleviating BID symptoms (Blom et al., 2012). In an online survey conducted on 54 BID individuals, Blom et al. (2012) observed that psychotherapy was often supportive and that antidepressants (but not antipsychotics) acted to mitigate the BID-related depressive symptoms. Still, both the psychotherapeutic and pharmacological treatments did not modulate the core symptoms of BID. The only treatment that, according to the authors, proved effective was actual amputation, as all those who underwent an amputation (7 out of the 54 examined cases) reported no longer experiencing BID symptoms. However, amputation of a healthy limb comes with certain health risks for the individual and constitutes a significant burden for the health system and society in general. The study of the neural correlates of BID can at the same time advance the understanding of BID and provide new knowledge on new practical rehabilitative treatments alternatives to amputation. For instance, by pinpointing altered neural patterns, it might be possible to tune these towards more neurotypical patterns through the application of non-invasive and non-harmful therapeutic intervention such as transcranial magnetic stimulation, Brain-Computer-Interfaces, or neurofeedback (Chakraborty et al., 2021; Ros et al., 2014).

Pioneering neuroimaging studies in this field focused on small samples of individuals with different limbs as targets of the amputation, and described circumscribed functional and structural alterations of the right superior parietal lobule (rSPL) and the left premotor cortex (lPMV; Hänggi et al., 2017; Hilti et al., 2013; McGeoch et al., 2011; van Dijk et al., 2013). We recently extended these findings in the so far most extensive neuroimaging BID study (Saetta et al., 2020). We gathered data on local structural changes and functional connectivity from 16 affected men with a homogenous manifestation of BID, i.e., the long-lasting and exclusive desire to have the left leg amputated. The results led to the first comprehensive model for the neural underpinnings of BID, suggesting i) reduced intrinsic functional connectivity of the primary sensorimotor area hosting the ‘to-be removed’ leg representations and the right paracentral lobule (rPCL, Saetta et al., 2020), suggesting that the feeling that a leg does not belong relates to a deficit in transferring information from the primary sensorimotor regions to higher-level cortical hubs for body processing; ii) anomalies of the lPMv (van Dijk et al., 2013; Blom et al., 2016; Saetta et al., 2020), which is thought to integrate the visual, motor, and tactile information about the affected limb into a coherent and unitary body representation (Ehrsson et al., 2005); iii) structural and functional abnormalities of the rSPL as a crucial neural correlate of body image, i.e., the conscious representation or visual template of one’s body size and shape (Hilti et al., 2013; McGeoch et al., 2011; Ramachandran et al., 2009; Saetta et al., 2020).

Outside of this network for sensorimotor and high-level bodily processing, a growing body of research suggests that the reward system and limbic circuitry may be altered in BID. Oddo-Sommerfeld et al., (2018) recently showed a heightened activity of the nucleus caudautus when individuals with BID looked at an image of themselves in the desired bodily state as compared to a control group. Moreover, the left orbito-frontal cortex (lOrbFG) as a key hub involved in both limbic and reward systems showed reduced intrinsic functional connectivity (Saetta et al., 2020). Other brain regions functionally altered in BID are the left superior and middle temporal gyri (lTG, Oddo-Sommerfeld et al., 2018; Saetta et al., 2020). These areas are often linked to the so-called mirror network. This network scaffolds the capacity to share others’ actions and emotions by their simulation through one’s own sensorimotor and affective systems (Gallese, 2006), and thus providing a neural basis for empathy (Bernhardt and Singer, 2012; Corradini and Antonietti, 2013; Gonzalez-Liencres et al., 2013; Pacella et al., 2017; Shamay-Tsoory, 2011).

However, as mentioned in a commentary by Longo (2020), even though our previous study (Saetta et al., 2020) had far-reaching implications for the understanding of BID, the analyses performed were primarily explorative. This calls for more hypothesis-driven neuroimaging studies spanning multiple methods of investigation to obtain a clearer picture of this multifaceted disorder. The current study takes a leap forward in this direction, identifying altered white matter structural connectivity (i.e., the anatomical connections between brain regions) in the same relatively large and homogenous sample of BID individuals. We quantified fractional anisotropy (FA) as a measure of structural connectivity derived from diffusion tensor imaging data (DTI). In brief, FA represents an elementary but robust tool to estimate anisotropic water diffusion within the cerebral white matter. Its values range between 0 – 1. Values approaching 1 reflect highly myelinated WM tracts as axonal cell membranes and myelin layers constrain the water diffusion along the axon’s length. In contrast, within tissues such as the cerebrospinal fluid, where the water diffusion is isotropic, unconstrained and random, values tend to be close to 0 (Pierpaoli et al., 1996; see below in the Discussion section for a more fine-graded picture of FA as drawn by high-resolution DTI methodology). Increasing evidence shows that the strength of the structural connectivity, as expressed by the FA measure, and functional connectivity positively and strongly correlate (Damoiseaux and Greicius, 2009; Greicius et al., 2009). Here we tested the hypothesis that reduced FA would be observed in the fronto-parietal hubs for which we previously found reduced intrinsic functional connectivity: the rPCL, the rSPL, the lOrbFG, the lTG. Additionally, given their implication in the limbic and reward systems (for a recent review see Palomero-Gallagher and Amunts, 2021), the right-hemisphere homologues of the lOrbFG were included.

## METHODS

### Participants

The BID sample is the same as that recruited for our previous study (Saetta et al., 2020), and consisted of sixteen men with the specific desire for amputation of the left leg (*Mean age* = 44.38, *SD* = 12.32, *Range* = 28-67; *Mean Formal Education in Years* = 16.06 *SD* = 2.28, *Range* = 13-18). Recruitment of BID individuals was performed through advertisements published on this website: http://www.biid-dach.org/. Neuroimaging data from 8 BID individuals were collected at the Department of Neuroradiology of the University Hospital of Zurich. Data from the other eight BID were collected at the Neuroradiology Department of the “ASST Grande Ospedale Metropolitano Niguarda” of Milan.

Sixteen healthy men functioned as controls (15/16 were the same recruited in the previous study, 1/16 was newly recruited as DTI data from one participant were not available). Controls were matched pair-wise to the BID individuals by age (*Mean Age* = 44.18, sd = 9.51, *Range* = 30-62 s, paired t-test: t(15) = 0.06, p = 0.95), years of formal education (*Mean Formal Education in Years* = 15.63, *SD* = 2.89, *Range* = 9.5-18, paired t-test: t(15) = 0.39, p = 0.70), and by scanner used for neuroimaging data acquisition (see below).

Informed consent was provided by participants prior to participation in the study, which had been approved by the Ethics Committee of the University Hospital of Zurich and by the Ethical Committee Milano Area C. The study was carried out according to the Helsinki declaration guidelines.

### Clinical assessments and questionnaires

As described in our previous studies (Hilti et al., 2013; Saetta et al., 2020), the presence of any neurological, psychiatric and neuropsychological disorders was formally assessed in all participants through standard examination and validated instruments. BID individuals additionally underwent the 2-h structured clinical interview for DSM-IV (Wittchen et al., 1997). Clinical examinations proved normal in both the control and the BID participants.

The clinical features of the desire for amputation were formally assessed through the 12-items Zurich Xenomelia Scale and its three sub-scores (Aoyama et al., 2012): “pure desire” (i.e., identity restoration as the primary goal of the desired amputation), “erotic attraction” (i.e., sexual preference towards amputated bodies), and “pretending” (i.e., the habit to simulate the desired body state by binding upon the undesired leg or using wheelchairs or crutches for locomotion). Each of the three sub-scores is computed by the mean of 4 items. For each item, answers are provided on a 6-points Likert-scale with 1 and 6 indicating, respectively, the least and greatest strong expression of the critical behavior or thought. Half of the items of each subscale are reverse-scored. The total score is the mean of all 12 items and represents the overall strength of the desire for amputation.

### Diffusion Weighted Images data acquisition

Diffusion Weighted Images (DWI) were acquired at two separate sites: the department of neuroradiology of the University Hospital of Zurich (Zurich dataset, n=16 (8 BID, 8 controls), and the Neuroradiology Department of the ‘ASST Grande Ospedale Metropolitano Niguarda’ of Milan (Milan dataset, n=16 (8 BID, 8 controls)). The Zurich dataset was acquired on a 3.0 Tesla Philips Achieva whole-body MRI scanner with an eight-channel head coil. Two identical diffusion-weighted sequences were acquired with a spatial resolution of 2 × 2 × 2 mm^3^ (matrix: 112 × 112 pixels, 75 slices in transverse plane). Diffusion was measured along 32 noncollinear directions (b = 1.000 s/mm^2^) preceded by a non-diffusion - weighted volume (reference volume, b = 0 s/mm^2^). The field of view was 224 × 224 mm2; echo time, 55.0 ms; repetition time, 13.010s; flip angle, 90°; SENSE factor, 2.1. Scan time was approximately 8 min 42 s per sequence.

The Milan dataset was acquired on a 1.5 Tesla General Electric (GE) Signa HD-XT scanner. Diffusion was measured upon 35 noncollinear directions (b =700 s/mm^2^) preceded by three non-diffusion-weighted volumes (reference volume, b = 0 s/mm^2^) with a spatial resolution of 1.094 × 1.094 × 4 mm^3^.

### DWI analysis

DWI data were processed using ExploreDTI (Leemans et al., 2009). Images were corrected for subject motion and eddy currents using the procedure described in Leemans and Jones (2009). Tensor estimation was then performed using the iteratively reweighted linear least-squares approach (Veraart et al., 2013). Whole-brain deterministic tractography was performed using DTI with a seed point resolution of 2 × 2 × 2 mm and Fractional Anisotropy (FA) threshold of 0.2. A restricted tractography analysis was performed subsequently to reconstruct streamlines passing through pairs of ROIs that form part of the Automated Anatomical Labelling (AAL) atlas (Rolls et al., 2015). By adopting a hypothesis-driven approach, we looked at reconstructed streamlines between selected ROIs and 90 regions of the AAL covering the whole cerebrum and subcortical structures excluding the cerebellum. These ROIs were: the right superior parietal lobule (AAL label: 60, Parietal_Sup_R), the right paracentral lobule (AAL label: 70, Paracentral_Lobule_R), the left middle temporal gyrus (AAL label: 85, Temporal_Mid_L), the left inferior temporal gyrus (AAL label: 89, Temporal_Inf_L), the left and right inferior frontal gyri, pars orbitalis (AAL labels: 11, Frontal_Inf_Oper_L; 12, Frontal_Inf_Oper_R); the left and right middle frontal gyri, pars orbitalis (AAL labels: 9, Frontal_Mid_Orb_L; 10, Frontal_Mid_Orb_R).

For each reconstructed streamline, Fractional Anisotropy (FA) was taken as the primary outcome measure. Additionally, the pairs of ROIs for which the number of participants presenting no streamlines was n > 8 were excluded from the analyses. This ensured that all the analyses included a minimum of n = 24 participants. Data were then manually checked to ensure an approximately equal distribution of missing values in the BID individuals and the control group. The FA matrices for each participant are available on the open science forum (OSF, https://osf.io/bdmsr/).

Group analyses were performed in R studio V. 1.3.1093. Data analysis scripts and their associated dataset are available on the OSF (https://osf.io/bdmsr/). Linear mixed models were fitted using R lme4 package (Bates et al., 2015) and estimated using REML and nloptwrap optimizer. The Wald approximation was used to compute the 95% Confidence Intervals (CIs) and p-values. A linear mixed model examined the FA values for each ROI pair as a function of the factor “Group” (BID individuals vs Controls). The factor “Scanner” defining the participants belonging to the Zurich or Milan dataset was modelled as a random factor to account for the data acquisition performed with different scanners (for a similar approach see Saetta et al., 2020). The tested statistical model was the following:

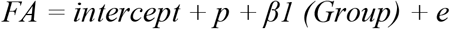

where “*β*_*x*_” stands for the estimated parameter, “*e”* stands for the residuals, and “*p*” represents the random intercept for Scanner.

The altered patterns of FA were correlated with the Zurich Xenomelia Scale scores through Spearman correlations, given the non-normality of data distribution.

## Results

### DWI analysis

Compared to controls, BID individuals showed significantly reduced FA values for the fibre bundles connecting the right superior parietal lobule and i) the right cuneus (*Mean FA BID =* 0.41, *Mean FA Controls =* 0.49, *b =* 0.072, *SE =* 0.012, *t(*27*) =* 5.707, *p <* 0.0001); ii) the right superior occipital gyrus (*Mean FA BID =* 0.42, *Mean FA Controls =* 0.46, *b =* 0.049, *SE =* 0.013, *t(*29*) =* 3.742, *p =* 0.0008); iii) right posterior cingulate gyrus (*Mean FA BID =* 0.47, *Mean FA Controls =* 0.50, *b =* 0.029, *SE =* 0.013, *t(24) =* 2.119, *p =* 0.045). Other ROI pairs with reduced FA values in BID were the following: the pars orbitalis of the middle frontal gyrus and the putamen in the right hemisphere (*Mean FA BID =* 0.43, *Mean FA Controls =* 0.45,*b =* 0.026, *SE =*0.01, *t(*27*) =* 2.543, *p =* 0.017), and the pars triangularis of the inferior frontal gyrus and the middle temporal gyrus in the left hemisphere (*Mean FA BID =* 0.42, *Mean FA Controls =* 0.44, *b =* 0.025, *SE =* 0.011, *t(*23*) =* 2.413, *p =* 0.024).

Conversely, BID individuals showed increased FA values for the fibre bundles connecting the right paracentral lobule and the right caudate (*Mean FA BID =* 0.43, *Mean FA Controls =* 0.3*9, b* = -0.034, *SE =* 0.012, *t(*22*) =* -2.812, *p =* 0.010); Results are displayed in Fig 1 and summarised in Table 1.

**Table 1.** Altered Fractional Anisotropy (FA) in BID compared with controls. Mean FA values and results of the linear mixed model procedure.

| White Matter Fiber Bundle | Mean FA<br>in BID | Mean FA<br>in Controls | b | Standard Error | t | df | p |
| --- | --- | --- | --- | --- | --- | --- | --- |
| **Reduced Fractional Anisotropy in BID** |  |  |  |  |  |  |  |
| Right Superior Parietal Lobule -<br>Right Cuneus | 0.41 | 0.49 | 0.072 | 0.012 | 5.707 | 27 | $p < 0.0001$ |
| Right Superior Parietal Lobule -<br>Right Superior Occipital Gyrus | 0.42 | 0.46 | 0.049 | 0.013 | 3.742 | 29 | $p = 0.0008$ |
| Right Superior Parietal Lobule -<br>Right Posterior Cingulum Gyrus | 0.47 | 0.50 | 0.029 | 0.013 | 2.119 | 24 | $p = 0.045$ |
| Right Middle Orbitofrontal Gyrus -<br>Right Putamen | 0.43 | 0.45 | 0.026 | 0.01 | 2.543 | 27 | $p = 0.017$ |
| Left Inferior Frontal Gyrus,<br>pars triangularis -<br>Left Middle Temporal Gyrus | 0.42 | 0.44 | 0.025 | 0.011 | 2.413 | 23 | $p = 0.024$ |
| **Increased Fractional Anisotropy in BID** |  |  |  |  |  |  |  |
| Right Paracentral Lobule -<br>Right Caudate Nucleus | 0.43 | 0.39 | -0.034 | 0.012 | -2.812 | 22 | $p = 0.010$ |

**Figure 1.**
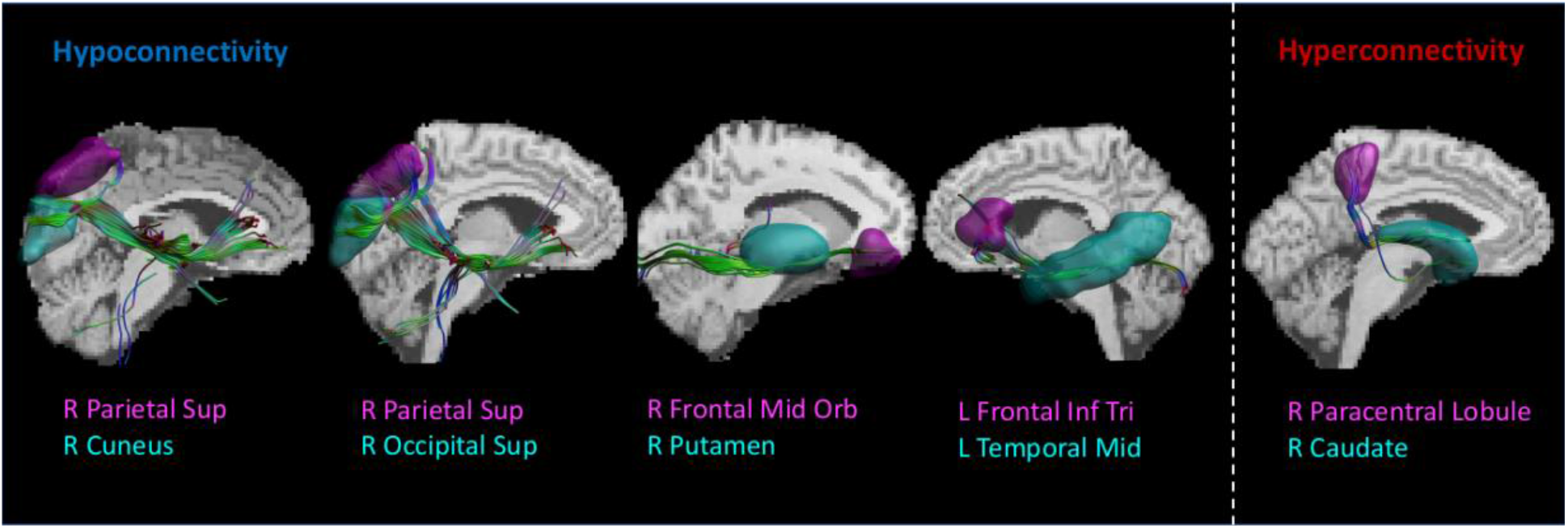
White matter pathways significantly altered in BID. Reconstructed streamlines superimposed on anatomical white matter template warped to individual subject data. Shown are tract visualisations for a representative healthy control participant,

### Correlations with the Zurich Xenomelia Scale

The mean sub-scores were: pure desire: Mean = 5.47, SD = 0.57, erotic attraction: Mean = 4.45, SD = 1.19, pretending: Mean = 4.24, SD = 0.66. Mean total score was 4.70, SD = 0.58. The FA values for the fibre bundles connecting the pars triangularis of the inferior frontal gyrus and the middle temporal gyrus in the left hemisphere negatively correlated with the strength of the desire for amputation (Spearman’s; r(11) = -0.63; p = 0.002) and the erotic attraction (Spearman’s correlation coefficient; r(11) = -0.58; p = 0.036) as assessed by the Zurich xenomelia scale. All the other correlations were not significant.

## Discussion

The results of the present DTI study, obtained in a homogenous and so far the most substantial sample of BID individuals (Longo, 2020), showed a reduced structural connectivity of i) the rSPL with the cuneus, the superior occipital and posterior cingulate gyri in the right hemisphere; ii) the rOFC with the putamen, iii) the triangular part of the left inferior frontal gyrus with the middle temporal gyrus. Increased structural connectivity was found between the rPCL and the right putamen. These results point to the presence of altered structural connectivity of cortical hubs previously found in the same sample to be functionally altered (Saetta et al., 2020).

In the discussion we address the different hubs in turn, beginning with those most classically associated with sensorimotor processing and bodily consciousness, before to move to those involved in other mental processes.

The rSPL showed reduced structural connectivity with visual retrosplenial regions such as the right cuneus, superior occipital and the posterior cingulate gyrus. This result is compatible with recent accounts regarding BID as a clinical condition that might ensue from a discrepancy between the body image represented in the rSPL (in BID individuals lacking a limb) and the actual “fully-limbed” functional body (McGeoch et al., 2011; Saetta et al., 2020). Accordingly, the binding up of the undesired leg frequently observed during pretending in BID individuals would temporarily solve this discrepancy and alleviate the desire through the adjustment of the visual information about the actual body to the visual aspects of the body image represented in the rSPL. This conceptual framework might be substantiated by the correlative evidence showing that the less concentration of gray matter in rSPL, the stronger an individual’s amputation desire and pretending behaviour (Saetta et al., 2020). In line with this, it has been shown that BID individuals experience a temporary relief from the desire for amputation, when the undesired leg is experimentally made to fade off from visual awareness (i.e., disappearing limb trick, (Stone et al., 2018) or while undergoing augmented or virtual reality to transiently adapt the appearance of illusory embodiment to the desired body opening the way for non-invasive therapeutics (Turbyne et al., 2021). Importantly, in line with this preliminary evidence suggesting that the mismatch between phenomenal and physical body would engage the visual representations, our findings show a specific pattern of reduced structural connectivity of the rSPL with visual processing brain areas.

Alongside this parieto-occipital network’s alteration, we found reduced structural connectivity between limbic system structures such as the rOFC and the putamen. These findings might be compatible with a compelling, yet speculative, model proposed by Ramachandran et al., (2009), according to which the body image in the rSPL would predispose an individual to be more sexually attracted by bodily shapes congruent with this template. The sexual preference would be mediated by the limbic and extrastriate areas, and the connection between them. While on a phenomenal level a body image lacking a limb as observed in BID individuals would determine a sexual preference to amputated bodies, on a neural level alteration in the connectivity between rSPL and visual and extrastriate areas might be further associated with alterations within the limbic system. In line with this model, in the current study we also found reduced FA in BID in the left uncinate fasciculus traditionally considered, by virtue of its location and connectivity (Von Der Heide et al., 2013) to be part both of the limbic (Hasan et al., 2009) as well as of the mirror systems (Wang et al., 2018), and, more specifically, of a fiber bundle connecting the left inferior frontal gyrus and the middle temporal lobe. Interestingly, while a growing body of research reveals a tight association between the integrity of the left uncinate fasciculus and individual levels of emotional empathy (Parkinson and Wheatley, 2014), alterations in empathic responsivity have been discussed as a social co-determinant of BID. Specifically, an excessive merging of BID individuals’ bodily self with that individuals with an amputation could arguably lead to an overidentification with amputated bodies, a process supposedly mediated by the mirror system (Brugger and Lenggenhager, 2014; Hilti et al., 2013; Macauda et al., 2017). Accordingly, introspective reports of BID individuals show specific differences in the quality and quantity of emotional experiences with others who have a disability during childhood (Money et al., 1977; Obernolte et al., 2015). A large number of BID individuals describe a “trigger event” for their desire for amputation, i.e. the sight of an amputee (Brugger, 2011; Hilti et al., 2013; Aoyama et al., 2012). They also report a significantly higher exposure to disabled people in their childhood environment (Obernolte et al., 2015) and eventually start to experience the kind of erotic attraction frequently accompanying BID (Money et al., 1977; Obernolte et al., 2015). One possible explanation for the link between empathic admiration and erotic attraction is tentatively offered in the psychodynamic perspective offered by Money et al. (1977), who suggested the existence of a “*non-erotic imagery of masculine overachievement as providing an erotic turn-on in their amputee fantasies. The over-achievement imagery primarily consisted of amputees overcoming the adversity of a handicap*” (p. 125). However, self-report reconstruction of early childhood memories should generally be treated with caution as they could represent an attempt of later rationalization (Gallo, 2010). Still, the negative correlation we found between reduced FA values of a left fronto-temporal bundle, the desire for amputation and, of special interest, also the erotic attraction, may potentially constitute the first neural correlate for the above-mentioned link.

Increased FA in BID was found between a cortical-striatal bundle connecting the rPCL and the right caudate nucleus. While the rPCL is involved in the leg sensorimotor processing of the affected limb, the latter region has been consistently found to be implicated in the reward system (review in Grahn et al., 2008). Increased FA in the right caudate and cortical regions has been observed in individuals with an obsessive-compulsive disorder (Lochner et al., 2012; Cannistraro et al., 2007; Fontenelle et al., 2009), a severe condition characterized by recurrent intrusive thoughts, ideas, and impulses also known as “obsessions”. We speculate that the desire for an amputation of one’s left leg, initially a consequence of a functional disconnection between rPCL and rSPL (Saetta et al., 2020), might develop in to obsessive features through continuous reinforcement induced by the hyperconnectivity of rSPL and the rightcaudate nucleus. Such assumption remains however speculative, and active-task fMRI studies are needed to test further test such hypothesis.

To summarize, the results of the previous and current studies suggest that BID might reflect multiple alterations of distinct but interconnected neural networks for body processing (including limb representation, multisensory integration, and body image), for emotional and social processing as well as of the reward system.

Our study comes with several limitations. First, the sample size is relatively small. This is due to the rarity and secrecy of the disorder. Only after a decade of cooperation between different international research centers has it been possible to put together a relatively substantial and homogenous database of individuals willing to participate in BID research. Second, and again relating to the challenges and timescale for data acquisition with this rare cohort, the DTI dataset was acquired between 2013 and 2018. Methodology for quantifying white matter microstructure has improved vastly in the intervening time period, meaning that FA is no longer considered the gold standard measure (Alexander et al., 2001; Jeurissen et al., 2013; Tuch et al., 2002). In particular, FA estimates are unreliable in regions that contain crossing fibres, such as the most lateral projections of the corpus callosum (Jeurissen et al., 2011; Ruddy et al., 2017) where low FA values may paradoxically reflect densely packed (but directionally incoherent) fibre pathways. This may raise questions regarding the validity of some of our findings using tractography from cortical regions of interest as we were unable to use alternative more high resolution approaches to measure fiber density, limited by the fairly low number of diffusion directions in which data was acquired. Finally, our empirical approach was deliberately limited to uncover the neural mechanisms of BID and relate them to individual phenomenal experience. A more comprehensive approach would consider all the possible nonlinear interactions between biological, psychological, and social factors of this aberrant condition. Only an integrative, cross-disciplinary view that includes the brain, the mind, and society as equally important levels of analysis will eventually lead to further essential advances in understanding a multifaceted disorder such as BID.

## Data Availability

Data analysis scripts and their associated dataset are available on the Open Science Forum, OSF (https://osf.io/bdmsr/).

## Acknowledgments

We thank the participants for their time and efforts as they traveled abroad to participate in the study. This work was supported by the Swiss National Science Foundation project (Grant Number: PP00P1_170511).

## Author contributions

Gsae wrote the manuscript. BLen, Pbru, Gbot conceptualized the project. BLen, PBru, GBot, acquired fundings. Gsae, Mgan, Lzap, Gsal, Msbe collected the neuroimaging data. Pbru, Gbot, Gsae, Mgan, Blen performed the clinical interview. Gsae, Krud performed data analyses. All authors revised and the manuscript and approved the final version.

